# Pathogenic *HFE* Variants and Evaluation for Hemochromatosis in Patients with Early-onset Atrial Fibrillation

**DOI:** 10.64898/2026.01.26.26344399

**Authors:** J. Michael Daw, Hollie L. Williams, Cassady J. Pelphrey, Dakota D. Grauherr, Diane M. Crawford, Dan M. Roden, Zachary T. Yoneda, Colleen T. Morton, M. Benjamin Shoemaker, J. Lukas Laws

**Author notes:** **Corresponding Author:** J. Lukas Laws, MD, Department of Medicine, Division of Cardiovascular Medicine, Vanderbilt University Medical Center, 1211 Medical Center Drive, Nashville, TN 37232, U.S.A. Authors contributed equally to the study (M. Benjamin Shoemaker, J. Lukas Laws). **Disclosures:** The remaining authors have nothing to disclose.

## Abstract

**Background:** Genetic testing is now recommended for select patients with early-onset atrial fibrillation (AF). Hemochromatosis is an autosomal recessive syndrome that occurs in patients who carry two pathogenic or likely-pathogenic (P/LP) variants in HFE. HFE is included on some genetic testing panels used for patients with AF. Hemochromatosis causes cardiomyopathy due to iron overload in the ventricle; however, it is unknown whether AF can be an early manifestation that is identified by genetic testing.

**Methods:** A total of 347 patients were referred to a dedicated AF precision medicine clinic. The clinical diagnostic evaluation included an H&P, 12-lead ECG, ambulatory ECG monitoring, and cardiac imaging (cardiac MRI and/or TTE). Genetic testing was performed using CLIA-approved laboratories: Labcorp/Invitae, GeneDx, or Vanderbilt University Medical Center. HFE was included on the cardiomyopathy panel used by 2 of the 3 laboratories.

**Results:** HFE was tested in 165 participants (median age 46 years [IQR 35-55], 115 [70%] male, 149 [90%] White). Six participants (4%) had two pathogenic variants in HFE. All of them were C282Y/H63D compound heterozygotes. Forty-one participants (25%) were heterozygous carriers of one pathogenic HFE variant. Among the 6 participants with 2 pathogenic HFE variants, the median ferritin level was 346 mcg/L [IQR 262, 496] (normal <300 mcg/L males, <200 mcg/L females). Three participants (50%) met laboratory criteria for iron overload. One individual had isolated ferritin elevation with normal transferrin saturation. All 6 underwent cardiac MRI as part of the genetic evaluation for early onset AF, and there was no evidence of cardiac siderosis based on cardiac T1 mapping median 990 ms [IQR 968-1024] (normal 960-1030 ms). Dedicated sequences to evaluate for iron overload demonstrated short hepatic T2* in one individual, indicating presence of hepatic iron overload (9 ms, normal >11.4 ms; liver iron concentration 3.4 mg/g, normal <2 mg/g). Three out of 6 participants were referred for a hematology evaluation and 2 out of 6 were started on therapeutic phlebotomy.

**Conclusion:** Genetic testing can identify patients with early-onset AF who are genetically susceptible to hemochromatosis, have evidence of iron overload, and receive early intervention with therapeutic phlebotomy. These results suggest HFE should be sequenced as part of genetic testing for early-onset AF, but larger sample sizes are needed to confirm these results.

---

Genetic testing is now recommended for select patients with early-onset atrial fibrillation (AF) to identify pathogenic variants that increase susceptibility to cardiomyopathies and arrhythmias.^1^ Some, but not all, cardiomyopathy and arrhythmia genetic testing panels include *HFE*, the homeostatic iron regulator gene associated with hemochromatosis. Cardiomyopathy can be a manifestation of hemochromatosis due to iron deposition in the myocardium. Hemochromatosis is an autosomal recessive syndrome that only occurs in patients who carry two pathogenic or likely-pathogenic (P/LP) variants in *HFE*. Two pathogenic variants, C282Y (c.845G>A) and H63D (c.187C>G), are found in up to 6% and 14% of the general population, respectively. *HFE* variants are therefore commonly reported, making it important for clinicians to interpret these results in patients with early-onset AF. We report the prevalence of *HFE* variants and results from the clinical evaluation for iron overload and hemochromatosis in a cohort of patients who underwent genetic testing for early-onset AF in a precision medicine clinic.

A total of 347 patients were referred for genetic evaluation of early-onset AF and prospectively enrolled in an IRB-approved registry. Clinical phenotyping included history and physical, standard 12-lead ECG, ambulatory ECG monitor, cardiac imaging (transthoracic echocardiogram and/or MRI), genetic counseling and genetic testing. Genetic testing used comprehensive cardiomyopathy/arrhythmia panels (ranging 138-175 genes) from one of three CLIA laboratories (Labcorp/Invitae, GeneDx, or Vanderbilt University Medical Center Clinical Genetic Testing Laboratory).

*HFE* was included on genetic testing panels from 2 of the 3 commercial laboratories and sequenced in 165 (48%) participants. Among participants sequenced for *HFE*, median age was 46 years (IQR 35-55), 115 (70%) were male, and 149 (90%) were White. None previously underwent testing for hemochromatosis. Forty-one participants (25%) carried a single heterozygous, pathogenic *HFE* variant (C282Y=13, H63D=28) requiring no additional workup. Six participants (4%) had two pathogenic variants in *HFE*; all six were C282Y/H63D compound heterozygotes. Iron studies in these participants showed a median ferritin level 346 mcg/L [IQR 262, 496] (normal <300 mcg/L males, <200 mcg/L females). Three participants (50%) met laboratory criteria for iron overload. A fourth individual had isolated ferritin elevation with normal transferrin saturation. All 6 underwent cardiac MRI demonstrating normal left ventricular function and no evidence of cardiac siderosis based on T1 mapping (median T1 time=990 ms [IQR 968-1024; normal range: 960-1030 ms). Dedicated sequences to evaluate tissue iron deposition demonstrated short hepatic T2* in one individual, indicating presence of hepatic iron overload (9 ms, normal >11.4 ms; liver iron concentration 3.4 mg/g, normal <2 mg/g). The 3 participants with evidence of serologic iron overload were referred for hematology consultation, and 2 started therapeutic phlebotomy based on their genetic predisposition and serologic values.

Overall, we found that 25% of patients tested for *HFE* were heterozygous carriers of one P/LP variant, and 6 (4%) carried two. This confirms previous findings that *HFE* mutations are common, especially among populations of northern European ancestry.

The penetrance of hemochromatosis varies by specific *HFE* genotype, which should be considered when interpreting genetic testing results. C282Y homozygosity carries the highest risk of iron overload (38-50%) and clinical hemochromatosis (10-33%).^2^ These individuals are recommended to undergo annual measurement of ferritin and transferrin saturation, with surveillance until age 55 if testing remains normal. C282Y/H63D heterozygotes are reported to have significantly lower risk of developing iron overload (3-24%), although hemochromatosis may develop in the presence of other risk factors such as fatty liver disease, diabetes, and alcohol use.^2-3^ Iron overload is less common although poorly characterized in H63D homozygotes. They should undergo screening, especially with a positive family history or symptoms of iron overload (most commonly liver disease and arthritis). Heterozygous carriers have equal risk to the general population and require no further evaluation.

Abnormal iron studies warrant hematology referral and investigation of alternative causes of hyperferritinemia. In our cohort, two compound heterozygotes were recommended to start routine phlebotomy by hematologists to lower the risk of progression to end-organ damage. Additionally, one patient was determined to have elevated ferritin in the setting of hepatitis B infection.

It is important to identify iron overload in patients at risk for hemochromatosis because early treatment and lifestyle modification reduces cardiovascular mortality.^4^ Historically, hemochromatosis is identified late in the course of disease when permanent organ damage has occurred. In this cohort, genetic testing identified patients with biallelic pathogenic *HFE* mutations and serum iron overload without cardiac siderosis or ventricular dysfunction, eligible for therapeutic phlebotomy. In addition to T1 mapping, dedicated T2* MRI sequences are necessary to detect and quantify tissue iron deposition in the liver and ventricle, and provides useful diagnostic information in early onset AF evaluation.^5^ Early detection enables cascade screening of relatives, biochemical surveillance, and targeted lifestyle interventions to reduce risk of cirrhosis by reduction in alcohol use and treatment of cardiometabolic disease. We recommend including *HFE* on genetic panels for early-onset AF and that cardiologists recognize these highly prevalent, but low penetrant *HFE* variants may prompt changes in management.

**Figure 1.**
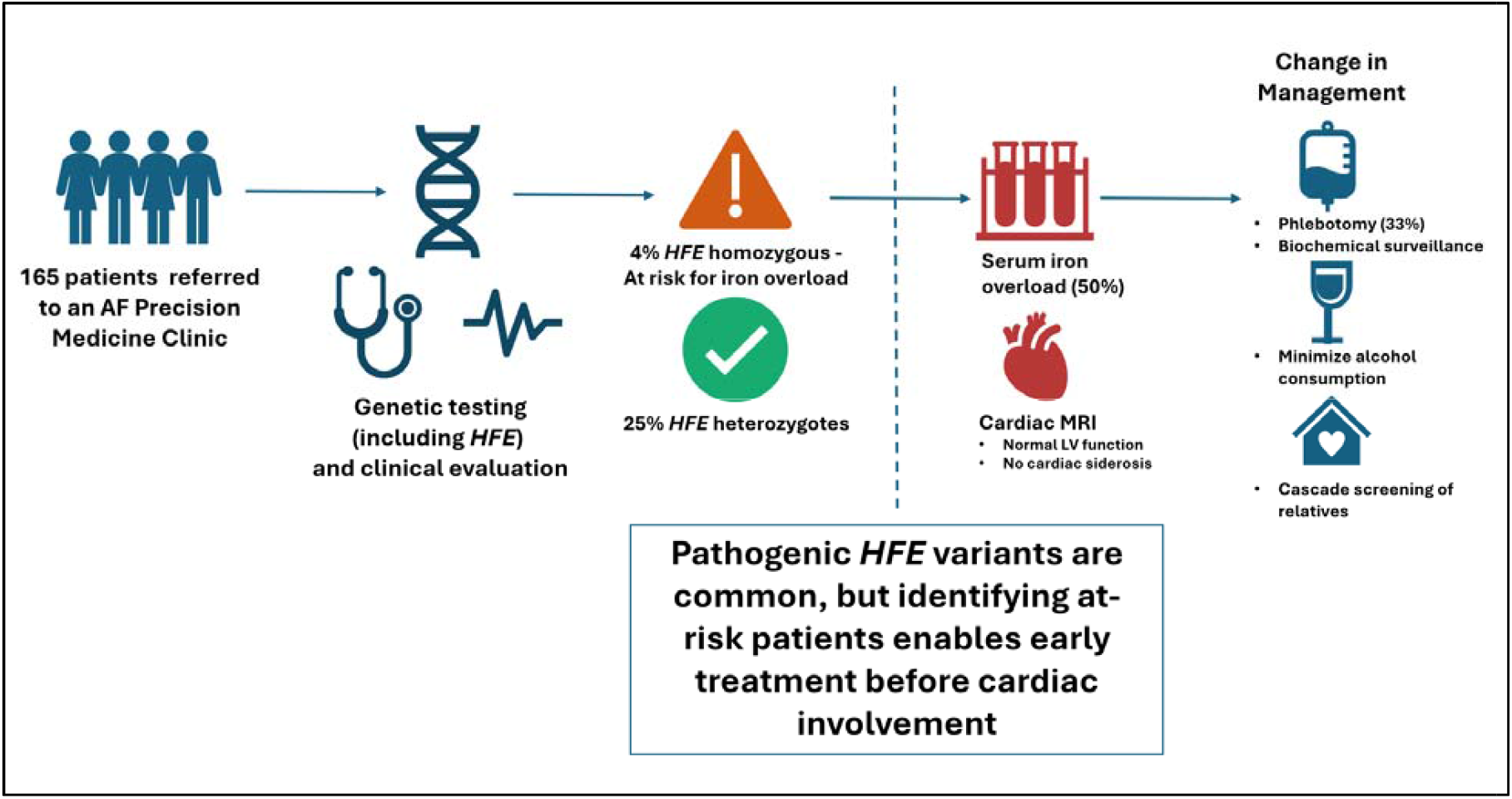
Evaluation and management of patients sequenced for *HFE*.

## Data Availability

All data produced in the present study are available upon reasonable request to the authors.

## Notes

### Competing Interest Statement

The authors have declared no competing interest.

### Funding Statement

This study did not receive any funding.

### Author Declarations

This study was approved by the Vanderbilt University Medical Center Institutional Review Board (IRB). All procedures performed in studies involving human participants were in accordance with the ethical standards of the institutional and/or national research committee and with the 1964 Helsinki Declaration and its later amendments or comparable ethical standards.

